# A service user evaluation of eConsult use by Defence Primary Healthcare Primary Care Clinicians using a mixed-method approach

**DOI:** 10.1101/2020.11.01.20220368

**Authors:** Antony S Willman

## Abstract

**Introduction:** eConsult has been recently introduced into Defence Primary Healthcare to allow patients electronic access to healthcare. Using mixed methods, this service evaluation sought the views of primary care clinicians using eConsult.

**Methods:** A two-phase sequential exploratory mixed-method approach was used. An inductive thematic analysis of feedback from primary care clinicians in the Salisbury Plain Area identified themes around eConsult use. These were used to construct an 18-item survey instrument. This was then distributed to primary care clinicians in Defence Primary Healthcare to assess the broader applicability of the themes.

**Results:** Four themes were identified: the impact on accessibility, the effects on working practices, the impact on the dynamics of the consultation and the effect of training and administrative support. eConsult did not save time for clinicians but was generally more convenient for patients. eConsult was often used in conjunction with telephone and face to face follow up, forming a ‘blended consult’. Accessibility was improved, but cultural factors may affect some patients engaging.

**Conclusions:** eConsult improves accessibility and can reduce telephone and face to face consultations but does not reduce workload. It should be used alongside conventional access methods, not instead of. It is useful for straightforward clinical and administrative problems but is less useful for more complex cases unless part of a ‘blended consult’. Future use could be modified to provide greater data gathering for occupational health and chronic disease monitoring and should be monitored to ensure it is inclusive of all demographic groups.

**KEY MESSAGES:** The increased accessibility to a clinician that eConsult offered for patients was positive.

There was no workload saving for clinicians using eConsult.

EConsult was often the first part of a ‘blended consult’ which subsequently involved telephone and face to face consults.

While the dynamics of the consultation were changed, this was generally perceived as positive.

EConsult should exist alongside current systems for accessing primary care.

The efficiency of the eConsult system could be improved with better administrative support and training.

## BACKGROUND

Remote consulting (RC) involves consultations with a doctor or other healthcare practitioner that can take place over the phone, online or via video link. Increased access to a primary care clinician (PCC) by RC and patients having the right to an online, or video consultation is a specific digital improvement enshrined in the NHS Long Term Plan ^(1)^. RC can provide patients with a convenient and easy way to ask for advice about straightforward problems, reducing their requirement for a Telephone (TC) or Face to Face consultation (FTFC) ^(2)^. This benefit is balanced by PCC concerns of increased workload, disadvantaging of certain groups, increased clinical risk and changing future practice without a rigorous assessment process ^(3)^. This reduction in FTF consultations and increased RC has accelerated because of the COVID-19 pandemic and the requirement to sustain primary care services while reducing risks to healthcare staff.

RC can either be a synchronous process (undertaken in real-time) such as via video teleconference (VTC) or TC, or it can be asynchronous as occurs with e-mail communication. An asynchronous tool already being used in primary care is eConsult (EC). This system was developed by practising GPs and piloted in 2014 ^(4)^. It provides amongst other things: self-care advice in the form of videos and symptom checkers; signposting to other services such as community pharmacy; a triage tool; the completion of a condition-based questionnaire requiring a time-limited response from the practice. EC aims to improve productivity and decrease inappropriate GP appointments. Early pilots did demonstrate access to GP services were enhanced. However, uptake was low, the self-help facility not always utilised, and the time saved in the context of reduced FTF consultations not always realised ^(5) (6)^. PCC satisfaction of EC improved if the patient problem was straightforward or modification of chronic disease management but less if the problem was more nuanced or complex ^(7)^.

Improvement in access, while beneficial and liked by patients, was seen by some staff as undermining other access methods such as telephone appointments with priority given to EC. Having to respond by the end of the next working day risked patients ‘gaming’ the system to prioritise their appointment. In summary, there are benefits to EC, but the introduction needs to be supported by staff engagement, protocols for the processes involved and regular audit ^(5)^.

Defence Primary Healthcare (DPHC) provides primary and occupational healthcare to Service Personnel (SP) in non-families practices and primary care to their dependents in families practices. EC had already been introduced as pilot schemes in some medical centres as part of Defence Healthcare Delivery Optimisation (DHDO). With the COVID-19 pandemic, a more rapid roll-out was undertaken. Within the area of Salisbury Plain Training Area (SPTA), three large and one small Defence Primary Healthcare (DPHC) group practices with a population of approximately 21000 patients have recently started using EC. It is crucial to understand how PCCs feel about EC as a new way of working and its impact on the relationship and delivery of care to patients. High-quality care should exploit technologies to reinforce solutions, not resolve non-existent problems. It is also essential to assess how staff and patients are safeguarded with this blended model of care. The current literature base is not well developed primarily due to EC being a recent concept. This evaluation aimed to add to the evidence base by exploring the views of PCCs recently introduced to EC in SPTA and test these views within the wider DPHC community who are already using EC. Due to the heterogeneous nature of RC (patient access, communication and consultation skills, IT literacy and workload issues, to name a few), a mixed-method approach was used.

## METHOD

To evaluate the views of PCCs using the EC system, a two-phase sequential exploratory mixed-method approach was used ^(8)^. An initial qualitative data collection from the SPTA practices was undertaken to construct an instrument for a more extensive quantitative and confirmatory survey of EC across DPHC. This second dataset was then integrated with the initial qualitative data to assess applicability of the themes as well as identify areas of convergence and divergence.

### Phase 1 - Qualitative data collection

#### Sampling and Data

There were three sources of data. The primary source was a Survey (SR) with responses collated from a simple post-consultation free text questionnaire designed using an online tool (Microsoft Forms) and completed by the PCC after an EC episode. The four SPTA DPHC Group Practices were invited to contribute to this via e-mail. The second source of data was mined during an online Journal Club (JC) where the paper by Brant et al. was discussed ^(3)^. This was recorded using Zoom with the permission of the participants, and the audio transcribed using an online transcription service. The third source of data was the transcript of a Skype Webinar Discussion (WD) involving EC stakeholders from London and South East region of DPHC. A list of EC problems, solutions and good practice points were discussed with themes captured by the author from the audio as well as text chat.

#### Data analysis

The qualitative survey was open for four weeks, with 48 SRs received from the practices from a total of 159 EC submissions (response rate 30%). PDFs of the SR, JC and WD dialogue were imported into Nvivo 12, where an inductive thematic analysis was performed. A mixture of ‘in vivo’ (verbatim) and transformation of comments created an initial framework of codes. The initial review of all three data sources provided 82 codes from 125 references which were grouped into 17 nodes in 4 themes. The codebook was shared with stakeholders in the SR, JC and WD groups who provided triangulation by confirming the nodes and themes were an accurate representation of the discussion

### Phase 2 - Quantitative data collection

#### Sampling and Data

The qualitative dataset was quantitised by using Likert scales to create a survey instrument for broader distribution amongst DPHC PCCs ^(9)^. Using the 17 nodes from phase 1, a series of statements about EC based were constructed and modified to reduce ambiguity ^(10)^. This produced an 18-point survey instrument. Six of the questions were reverse worded to try and reduce acquiescence bias ^(11)^. The Likert scale was scored between 0 (disagree) and 10 (agree) with 5 as neutral (neither agree nor disagree). Three additional questions were added asking about role, families or non-families practice, and a free text box was added for feedback on EC. REDCap was used to design the survey and a hyperlink distributed to all PCCs within DPHC via e-mail with explanatory text. 134 responses were received over 4 weeks. At the time of the survey, the EC mailbox list had 433 PCC users giving a response rate of 31%.

#### Data analysis

Data was imported, cleansed and analysed using Microsoft Excel. Given the ordinal nature of data from Likert scales, medians and interquartile ranges (IQR) were calculated to assess the degree of agreement by the respondents to the survey statements. Agreement with positively and disagreement with negatively worded statements would support applicability across the respondents.

## RESULTS

Of the 134 PCC responses, 60 (45%) worked in a families practice and 74 (55%) in a non-families practice. 117 (87%) were GPs with the remaining 17 (13%) being composed of other PCCs. Data from phase two is represented in Figure 1. There was agreement with all of the positively worded statements and disagreement with three of the negatively worded statements. Three were neutral (‘EC saves time for the clinician compared to telephone or face to face consultations’, ‘EC saves time for the clinician compared to telephone or face to face consultations’, ‘EC is a more convenient way for a clinician to deal with patient problems’) but with the 1^st^ quartile range weighted towards disagreement.

### Impact on accessibility

Text quotes were taken from both datasets.

It was broadly recognised that EC improved accessibility for patients allowing them to undertake a consultation at a time and place of their choosing; a significant benefit for patients. Timely contact, ease of booking an appointment and convenience were all positive factors. The comfort of asking about personal or embarrassing problems was also felt to be increased.

*‘Too early in eConsult implementation across DPHC to draw any firm conclusions. Initial feedback is that patients like it and adds to accessibility of healthcare. From a GP perspective, it has probably increased workload due to the extra admin associated and duplicate appts (e*.*g. cases still needing a phone call or F2F)’*. GP203

Increased accessibility did have drawbacks. The ease of access was reflected in some references as potentially increasing workload without the ‘safety net’ of negotiating normal gatekeeping channels.

*‘The immediate access to clinicians without limitation at any time of day will very likely lead to decreased usage of other services (pharmacy, Highstreet opticians etc) and there may be value in a first order screening mechanism to redirect inappropriate/ trivial traffic’*. GPST162

Barriers to the uptake of EC identified included Chain of Command buy-in and transcultural issues; the latter a particular problem with the Service Nepalese community. Advertising, using a link in an e-mail signature block and identifying patient advocates within certain service communities may improve this.

*‘EC is VERY population dependent - the uptake in a Gurkha unit has been almost non-existent, despite regular advertisement and encouragement. The language barrier is, I suspect, an issue’*. GP158

EC may have a role as a triage tool but needs to have a secure safety net and should co-exist alongside existing access systems. For straightforward issues, EC provided a good way for the patient to get a response quickly with or without further contact. The increased accessibility of EC does increase workload, both administrative and clinical, with several comments about poor manning.

*‘I am not convinced it saves time in small practices without dedicated eConsult admin/nursing support. I can see the benefit in a big, well-staffed practice if dedicated admin support can be allocated to triage. eConsult is useful for repeat prescription requests and admin queries that can be answered within eConsult’*. GP200

### Effects on working practices

Another benefit was the quality of the completed EC and any attached images. If the history provided enough detail and was combined with a good quality image, diagnoses could be based on the EC alone for certain skin conditions. The requirement to review the PHR meant this process was time equivalent to a FTFC, not time-saving.

*‘It is useful for skin conditions when avoiding face to face consultations. It would work well as a triage system. It lacks the personal interaction between doctor and patient, therefore lacking the ability to reassure patients, to build rapport. Appointment times need to be longer for e-consult because further investigation into the patient’s medical records is needed to confirm PMH/meds etc*.*’* – GP42

Processes such as occupational health requests and repeat prescriptions were also areas respondents felt EC was useful for. More efficiency in the appointment system was cited as being beneficial, with telephone calls being replaced by EC. This was both for the clinician responding to straightforward requests, and the patient having to phone in for something.

*‘l’d agree-has saved me many appointments and can sort scripts quickly and send message to patients when to collect. Previously many would book telephone appointment which took 15min slot. I can see how eConsult can be useful for this type of problem*.*’* - SR

However, there was a consensus that there was no overall time saving by using EC. Paperwork, pdf uploading, and repeat consulting all meant that while some consults were simple and straightforward, the workload balanced out.

*‘Took longer for both of us than if he had simply booked a F2F consultation…still have to review patient and produces extra paperwork in the process*.*’* - SR

Across all datasets was the concern that patients were adapting to the system, using EC to bypass the normal processes for booking appointments and getting preferential treatment. Deliberate submission of clinical issues via the administrative route bypassing the symptom checker was also mentioned. This invariably resulted in a TC due to the lack of clinical detail on the EC.

*‘Yes, but they don’t speak to reception, they get a GP to call them straight back, same day. Of course, they love it. That’s what’s happening. It’s just a way of avoiding booking appointments*.*’* – JC

### Impact on the dynamics of the consultation

There were mixed views as to how the dynamics of the consultation were changed. The lack of non-verbal communication and the human factor was felt to make EC appropriate for certain transactional types of consultation but not more complex ones.

*‘Patients are frustrated with the time required to complete the form and see it as a barrier to discussion with a clinician…blended consultations are a good way of added efficiency, but this software package is not right for the military med centre or palatable to our population*.*’* GP130

Acceptance that EC was part of a ‘blended’ consult was supported. Several respondents felt this was efficient, was convenient for the patient and was analogous to sequential consultations held commonly in primary care. Other respondents felt the role of EC was purely as a process to access a PCC and should not be considered part of the consultation.

*‘Better use of eC for chronic disease management is the way forward, eC can be used to data gather before we conduct the review F2F or over the phone. I feel that many medication reviews could be conducted in this way also by sending the patient a review template and asking them to complete it before we will renew their prescription’*. ANP125

*‘I see eConsult as a digital tool to access the practice. I don’t see it as part of a consultation, and I am not sure that it is meant to be. It is just another way for the patients to get in touch. It is not a replacement for a consultation’*. GP7

### The effect of training and administrative support

There was scope for improving the training of both clinical and non-clinical staff as well as the provision of adequate administrative support. The EC required effective triage to ensure it reaches the most appropriate person (administrative or clinical).

*‘Admin staff are not clinical admin staff; it takes as long if not longer for a standard admin to signpost a request. There needs to be a lot more training*.*’* GP204

There were several mentions of EC being used to capture lifestyle data (e.g. smoking status) for entry onto the Patient Health Record (PHR).

## DISCUSSION

Synthesis of the quantitative data provided by the survey demonstrated the majority of statements formulated from the qualitative phase were applicable across DPHC. Positive features of EC included improving accessibility, teledermatology, repeat prescription requests, administrative requests and as a tool to advise on non-complex clinical problems. The self-help feature was useful. Some respondents felt accessibility increased workload, but this seemed to be as a consequence of lack of training of non-clinical administrative staff, manning and poor triage of EC. This interface, where EC arrives in the practice, is crucial to future success. It involves defining the roles of clinical and non-clinical HCWs with the provision of appropriate training. In a DPHC Practice, this will involve administrative staff and Combat Medics (CMT). It is akin to a triage process, with the need to differentiate safely between the urgency and types of problems ^(12)^. This safety aspect was one of the less positive aspects of EC. Others included questioning the reasons for the introduction of EC and the minimal impact on workload. These concerns are not new ^(3)^. While there may be efficiencies and improvements in productivity, the universal view in both datasets was that EC should exist alongside current access systems, not instead of.

The dynamics and transactional nature of the consultation and the role of EC produced interesting results worth exploring. The idea of the ‘blended consult’ was supported by the majority of respondents suggesting EC was more than just a triage tool. Conversion rates to either FTFC or TC were not evaluated in this study but previously have been shown to approach 70% ^(6)^. This does not necessarily mean EC is inefficient but more of a precursor to a subsequent and more targeted consultation. For chronic disease for example, promoting aspects such as self-management are important patient enablers ^(13)^. Achieving this involves psychosocial enquiry, acknowledging non-verbal cues and exploring ‘ideas, concerns and expectations’, all of which are associated with increased quality of, and satisfaction with the consultation ^(14) (15)^. These tasks are unlikely to be achieved by RC alone. Therefore, the role of EC as a data-gathering tool may be the limit of what it can add to the ‘blended consult’ until use of the system matures.

The services offered by DPHC include occupational healthcare (OH), as well as close liaison with the Chain of Command (CoC), the employer, making it very different to most NHS settings. DPHC is a more patient-facing and probably patient focussed system because of this. Respondents reported that straightforward OH issues can be resolved by EC, but it was less suited for more complex cases. This is also the case with the holistic and pastoral care to SP that DPHC offers. EC may offer an entry point to a PCC but dealing with an ‘unhappy’ SP, and the importance of both verbal and non-verbal communication, is best achieved by FTFC ^(16)^. The flexibility of the open access that DPHC offers to the CoC for urgent assessment of SP must not be lost in the desire to expand EC use. Another area of interest is the difference between DPHC users and NHS demographics. Edwards et al. noted the poor uptake by 18-24-year-olds which represents a sizeable cohort of Service Personnel (SP) ^(6)^. There is likely to be a higher number of musculoskeletal, sexual and mental health presentations, some of which SP may be reluctant to present with. The increased accessibility and ability to contact a PCC at a convenient time offered by EC may well prove to be a positive enabler. This needs to be balanced against the possible disadvantage for Foreign and Commonwealth soldiers, especially the Nepalese community, highlighted by several respondents. The use of EC champions in both the service user community and Chain of Command may improve uptake.

### Limitations of this study

This study looked purely at the views of PCCs within DPHC. Views of administrative staff and military paramedics were gathered but not included in this analysis. This represents an important cohort and needs to be explored. Views of patients have not been included in this analysis. This aspect, especially the patient journey from the decision to initiate an EC to the subsequent outcome, should be a matter of priority for DPHC. Extrapolating how this journey differs between different cultural and demographic subsets will provide valuable insight into how EC can be improved.

## CONCLUSION

EC is a useful addition to current DPHC primary care services. It should be used alongside conventional methods of access but promoted for specific uses, e.g. simple clinical and occupational health requests, administrative requests, repeat prescribing and teledermatology. It can form part of a ‘blended consult’ where it is the first point of contact for a sequence of consults that may involve further ECs, TCs or FTFCs. Safe and effective triaging to the most appropriate HCW to deal with an EC request is crucial and there is a requirement to ensure both initial and ongoing training. EC can improve patient access but should not be seen as a panacea - simple changes such as additional phone lines and ensuring adequate staffing are equally as important. Workload has been unchanged by its introduction. There should be a process of ongoing monitoring to assess the uptake in groups who are less likely to engage with EC, e.g. young males and certain ethnic groups.

## Data Availability

All data is in the article.

