## Supplemental appendix A for "A service user evaluation of eConsult use by Defence Primary Healthcare Primary Care Clinicians using a mixed-method approach"

| Theme | eConsult Journal Club | eConsult Survey Response | eConsult Webinar Comment | Total |
|---|---|---|---|---|
| | References | | | |
| 1 : Impact on accessibility | 9 | 33 | 9 | 51 |
| Nodes | Barriers to access exist but can be overcome with measures including advertising and Chain of Command engagement<br>eC can be more convenient for the patient as well as for the clinician.<br>eC can improve access for patients and reduce the need for face to face consultations but should not be only way of accessing a clinician.<br>eC can increase the efficiency of consulting, especially with straightforward problems, but can also help with difficult or embarrassing issues.<br>eC may actually increase workload due to increased accessibility.<br>eC use as a triage tool can improve efficiency but it should have a safety net, so patients seek help sooner if necessary. | | | |
| 2 : Positive and negative effects on workflow and working practices | 6 | 30 | 7 | 43 |
| Nodes | Ability to attach a photograph and share it securely is beneficial and helps the diagnosis.<br>Certain requests such as repeat prescribing can be facilitated by using eC.<br>eC does not save time for the clinician and can be a lengthy process for the patient.<br>eC may allow patients to bypass normal processes for booking appointments.<br>eC may provide efficiencies for the administrative team, for example by capturing up to date contact details and occupational health data.<br>The health ecomonics of eC need evaluating as it may prove more expensive than current practice. | | | |
| 3 : The impact on the dynamics of the consultation | 0 | 19 | 1 | 20 |
| Nodes | eC may be improved if there was limited two-way dialogue or if previous patient history could be incorporated into a new consult<br>The limitations of eC need to be recognised especially with complex problems, human interaction and non-verbal communication<br>eC is an efficient way to gather information and data from the patient which can be followed up by a phone call or response, making it a 'blended' type consultation | | | |
| 4 : The effect of training and administrative support | 3 | 6 | 2 | 11 |
| Nodes | There may be quicker and more efficient systems available less vulnerable to the IT infrastructure<br>Training and support for administrative staff will allow eC to work more efficiently | | | |
| Total | 18 | 88 | 19 | 125 |
