## Supplemental appendix B for "A service user evaluation of eConsult use by Defence Primary Healthcare Primary Care Clinicians using a mixed-method approach"

### eConsult Survey

Thank you for completing this survey

There are a couple of demographic questions at the beginning followed by a series of 18 statements which were identified from a smaller qualitative pilot. Please click or press on the Likert scale first then adjust to reflect how much you agree or disagree. The scale is graduated from 0 to 100 with 0 being disagree, 50 neutral and 100 agree.

The number of questions has been kept as low as possible to try and limit the statements to one theme only. If you are not sure or don't want to answer, please just leave the slider at 50 or 'neutral'.

All responses are anonymised so feel free to be as honest as you want. The responses can be based on your own experiences, those of your colleagues or within your practice.

There is a free text box at the end to add any comments about eConsult you feel are important. This is optional.

All data is kept on secure REDCap servers within the University of Birmingham.

eConsult is abbreviated to eC in the statements.

1) What is your role?

◯ Military GP ◯ Civilian Medical Practitioner ◯ Advanced Nurse Practitioner ◯ GPST ◯ GDMO or equivalent ◯ Other

2) Is your Practice a Families Practice?

◯ Yes ◯ No

#### EConsult and the Impact on accessibility

3) Barriers in accessing eConsult (eC) exist but can be overcome with measures including advertising and Chain of Command engagement

Disagree Neutral Agree

*(Place a mark on the scale above)*

4) eC is a more convenient way for the patient to access healthcare

Disagree Neutral Agree

*(Place a mark on the scale above)*

5) eC can reduce the need for telephone or face to face consultations

Disagree Neutral Agree

*(Place a mark on the scale above)*

6) eC can increase the efficiency of consulting when dealing with straightforward problems

Disagree Neutral Agree

*(Place a mark on the scale above)*

7) The increased accessibility provided to the patient by eC decreases clinician workload

Disagree Neutral Agree

*(Place a mark on the scale above)*

8) eC use as a triage tool to access primary care can improve efficiency and should be the main way to access a clinician

Disagree Neutral Agree

*(Place a mark on the scale above)*

#### EConsult and the positive and negative effects on workflow and working practices

9) The ability to attach a photograph to eC and share it securely is beneficial and helps the diagnosis

Disagree Neutral Agree

(Place a mark on the scale above)

10) Certain requests such as repeat prescribing can be facilitated by using eC

Disagree Neutral Agree

(Place a mark on the scale above)

11) eC saves time for the clinician compared to telephone or face to face consultations.

Disagree Neutral Agree

(Place a mark on the scale above)

12) eC may allow patients to gain advantage in accessing a clinician by bypassing normal processes for booking appointments

Disagree Neutral Agree

(Place a mark on the scale above)

13) eC can provide efficiencies for the administrative team, for example by capturing up to date contact details and occupational health data

Disagree Neutral Agree

(Place a mark on the scale above)

14) The health economics of eC need evaluating as it may prove more expensive than current practice models such as telephone consulting

Disagree Neutral Agree

(Place a mark on the scale above)

#### EConsult and the impact on the dynamics of the consultation

15) eC can gather information and data from the patient which can be followed up by a phone call or face to face appointment as part of a 'blended consultation'

Disagree Neutral Agree

(Place a mark on the scale above)

16) eC is a good system to use to deal with more complex problems

Disagree Neutral Agree

(Place a mark on the scale above)

17) The 'blended consultation' is a feasible future model for primary care

Disagree Neutral Agree

(Place a mark on the scale above)

18) eC is a more convenient way for a clinician to deal with patient problems

Disagree Neutral Agree

(Place a mark on the scale above)

#### The effect of training and administrative support on eConsult utilisation

19) There may be quicker and more efficient alternatives to eC which are less vulnerable to the MOD IT infrastructure

Disagree Neutral Agree

*(Place a mark on the scale above)*

20) Training and support for administrative staff will allow eC to work more efficiently

Disagree Neutral Agree

*(Place a mark on the scale above)*

21) Please comment on any aspects of eC you think are important and not captured in the above statements

__________________________________________
