## Supplemental Figure 1 for "A service user evaluation of eConsult use by Defence Primary Healthcare Primary Care Clinicians using a mixed-method approach"

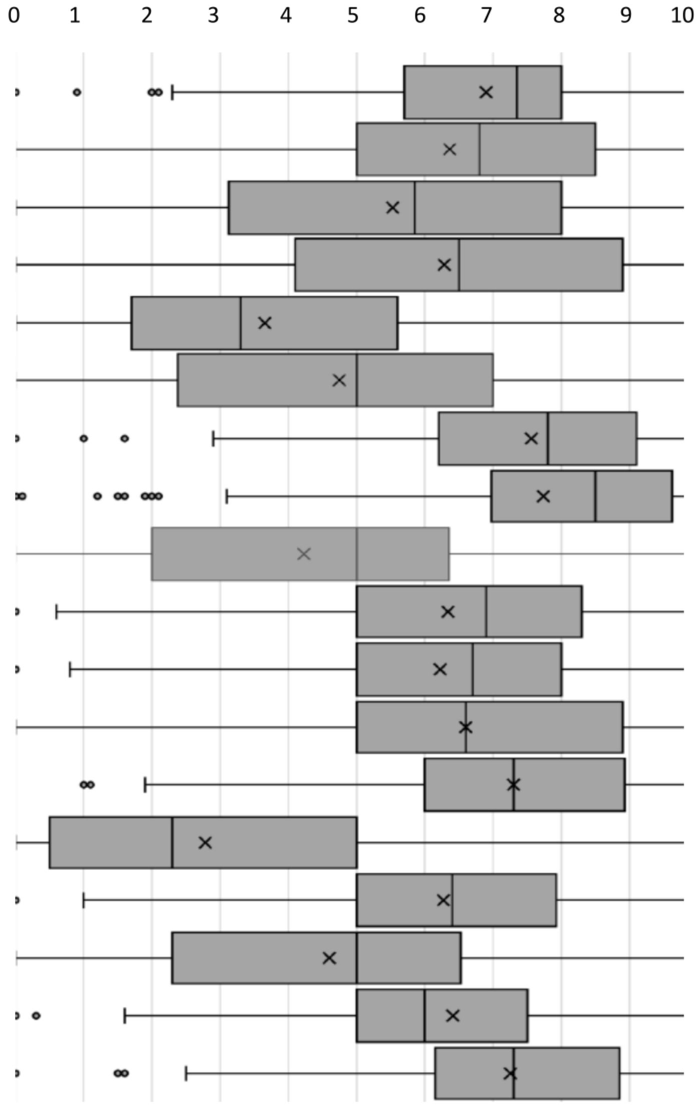


| Survey Statement | Median (IQR) |
|---|---|
| Barriers to access exist but can be overcome with measures including advertising and Chain of Command engagement. | 7.3 (5.7-8) |
| EC is a more convenient way for the patient to access healthcare. | **6.7 (5-8.25)** |
| **R -** EC can reduce the need for telephone or face to face consultations. | **5.8 (3.05-7.95)** |
| EC can increase the efficiency of consulting when dealing with straightforward problems. | **6.5 (4.03-8.75)** |
| **R -** The increased accessibility provided to the patient by EC decreases clinician workload. | **3.05 (1.71-5.33)** |
| **R -** EC use as a triage tool to access primary care can improve efficiency and should be the main way to access a clinician. | **5 (2.4-7)** |
| The ability to attach a photograph to EC and share it securely is beneficial and helps the diagnosis. | **7.65 (6.13-9)** |
| Certain requests such as repeat prescribing can be facilitated by using EC. | **8.3 (6.9-9.5)** |
| **R -** EC saves time for the clinician compared to telephone or face to face consultations | **4.7 (2-6.13)** |
| EC may allow patients to gain advantage in accessing a clinician by bypassing normal processes for booking appointments. | **6.85 (5-8.2)** |
| EC can provide efficiencies for the administrative team, for example by capturing up to date contact details and occupational health data. | **6.6 (5-7.9)** |
| The health economics of EC need evaluating as it may prove more expensive than current practice models such as telephone consulting. | **6.5 (5-8.8)** |
| EC can gather information and data from the patient which can be followed up by a phone call or face to face appointment as part of a 'blended consultation'. | **7.2 (6-8.9)** |
| **R -** EC is a good system to use to deal with more complex problems. | **2.3 (0.5-5)** |
| The 'blended consultation' is a feasible future model for primary care. | **6.4 (5-7.9)** |
| **R -** EC is a more convenient way for a clinician to deal with patient problems. | **5 (2.3-6.4)** |
| There may be quicker and more efficient alternatives to EC which are less vulnerable to IT infrastructure. | **6 (5-7.5)** |
| Training and support for administrative staff will allow EC to work more efficiently. | 7.25 (6.1-8.73) |

Boxes represent quartile ranges either side of median. X = Calculated mean. ● = Outliers (>1.5x IQR below 1st quartile). Whiskers represent range of values. **R** – reverse worded question.
